# Multi-Model Comparison of Deep and Classical Architectures for ECG Diagnostic Superclass Classification on PTB-XL

**DOI:** 10.64898/2026.10.25.26366433

**Authors:** Vivaan Bhatt, Jordan B. Strom, Francisco Lopez-Jimenez, Demilade A. Adedinsewo, Venkatesh L. Murthy, Alessandro Hammond, Thoriso Diale, Fu Siong Ng, Martin R. Cowie

**Affiliations:** Northville High School, Northville, MI, USA; Harvard University, Cambridge, MA, USA; Beth Israel Deaconess Medical Center and Harvard Medical School, Boston, MA, USA; Department of Cardiovascular Diseases, Mayo Clinic, Rochester, MN, USA; Department of Cardiovascular Medicine, Mayo Clinic, Jacksonville, FL, USA; University of Michigan, Ann Arbor, MI, USA; Score Academy, Pretoria, South Africa; Imperial College London, London, UK

**Keywords:** Electrocardiogram (ECG) classification, PTB-XL, deep learning, state space models, Mamba, MiniRocket, LightGBM, ensemble learning

## Abstract

**Objective:** It remains unclear which architecture best sorts 12-lead electrocardiograms into diagnostic superclasses, and whether added model complexity pays off.

**Methods:** Using 21,375 PTB-XL records under one patient-level split, we compared a Mamba-style state space model, a residual convolutional network, a bidirectional long short-term memory network, a convolutional-recurrent network with attention, soft-voting ensembles of these networks, and MiniRocket features with LightGBM. Each record took the superclass of its highest-likelihood diagnostic code, and we scored saved predictions with paired patient-level bootstrap intervals.

**Results:** No model clearly outperformed MiniRocket with LightGBM, which needs no graphics processor. The ensemble of the three deep models with the highest validation macro F1 score reached 0.656, against 0.645 for MiniRocket with LightGBM (difference +0.011, 95% confidence interval −0.007 to +0.031), and an ensemble of all four deep models reached 0.657. An exploratory ensemble, whose members we chose after seeing test results, reached 0.665 and was the only ensemble whose interval excluded zero (+0.021, 0.002 to 0.040); we regard that margin as optimistic. Hypertrophy was the hardest class (F1 0.33–0.41), and differences in its recall mostly reflected threshold placement.

**Conclusions:** Under one shared split, deep ensembles added little over a classical pipeline.

**Significance:** These results give biomedical informatics researchers a statistically grounded reference for judging whether complex models justify their cost on this benchmark. With one dataset, one split, one training seed per model, and no external validation, they do not show readiness for clinical use.

## I. Introduction

Cardiovascular disease is still the leading cause of death worldwide [1], and the 12-lead ECG is one of the most widely available ways to detect it. Software that reads ECGs already exists, but assigning a recording to a diagnostic category is still hard. The abnormalities can be subtle, several conditions often appear in the same patient, and the categories are heavily imbalanced. Deep learning has even found signals that clinicians cannot read from the tracing, such as atrial fibrillation in ECGs recorded during sinus rhythm [2]. Systematic reviews cover deep learning for ECG data in much more depth [3], [4].

In this paper we compare deep learning and classical approaches on one task. The task is to assign each 12-lead recording in the public PTB-XL dataset to one of five diagnostic superclasses. These are conduction disturbance (CD), hypertrophy (HYP, which in PTB-XL covers left and right ventricular hypertrophy, atrial overload or enlargement, and septal hypertrophy), myocardial infarction (MI), normal ECG (NORM), and ST/T change (STTC). Convolutional networks have generally done well on this kind of task, and recurrent models suit the sequential nature of the signal. Selective state space models (SSMs) are a newer option for long sequences, and MiniRocket, a random convolutional feature transform, offers an alternative that does not use deep learning.

The six approaches are a Mamba-style SSM (M1), a 1D residual CNN (M2), a BiLSTM (M3), a CRNN with attention (M4), a soft-voting ensemble of M1, M3, and M4 (M5), and a MiniRocket plus LightGBM pipeline (M6). M5 combines the three deep models with the highest validation macro F1. We also report two other ensembles as extra checks. One averages all four deep models (M5A). The other is an exploratory ensemble of M2, M3, and M4 (M5T), whose members we chose after seeing test results. Every model with learnable parameters was trained from scratch, and all models were tested on the same 3,182 records, which share no patients with the training or validation data. Besides argmax macro F1, we report AUROC, AUPRC, per-class results, and calibration, and we compare pairs of models with paired, patient-level statistics. Our main finding is that no model was clearly better than MiniRocket+LightGBM. The ensemble chosen on validation was ahead of the Mamba-style SSM and the CRNN, but it was not detectably different from the ResNet, the BiLSTM, or MiniRocket+LightGBM. Only the test-selected ensemble M5T had an interval over MiniRocket+LightGBM that excluded zero, and that margin is probably optimistic.

## II. Literature Review

Wagner et al. released PTB-XL as a large, free, public 12-lead ECG dataset for reproducible machine learning work, with SCP-ECG diagnostic codes mapped to broader superclasses [5]. It is now a common benchmark for deep architectures in this area. Nonaka and Seita compared many deep architectures on PTB-XL and two other 12-lead datasets and found that a standard ResNet-18 performed consistently well [6]. The PhysioNet/Computing in Cardiology Challenge 2020 made multi-source 12-lead classification a shared task [7]. Convolutional networks, including residual ones [8], are standard benchmarks for this task [9], and their local filters suit features such as the QRS complex. Recurrent models built on LSTM units [10], and convolutional-recurrent hybrids, instead capture features that spread across the full 10-second, 12-lead recording.

State space models are a fairly recent alternative to conventional sequence models. Mamba makes the state-space parameters depend on the input and adds a hardware-aware implementation. This gives linear-time modeling of long sequences [11], which suits ECGs with thousands of samples. SSMs are not new to ECG work, though. Mehari and Strodthoff applied S4 models [12] to PTB-XL and reported a macro AUC of 0.942 on the 71-label task, above their previous best [13]. A Mamba-based classifier has also been tested on PTB-XL [14]. Our contribution is narrower. We compare a pure-PyTorch Mamba-style model with CNN, recurrent, and classical baselines on one split, without the original fused CUDA kernel.

Classical methods also stay competitive in many settings. ROCKET and its faster, nearly deterministic variant MiniRocket use random convolutional kernels to extract fixed features that do not depend on labels and can feed any down-stream classifier [15], [16]. LightGBM, a gradient-boosted tree framework, handles such features efficiently [17]. Soft-voting ensembles tend to help when the errors of the members are not strongly correlated [18].

Published PTB-XL numbers are hard to compare directly with ours for several reasons. Strodthoff et al. [9] treat the superdiagnostic task as multi-label and report macro AUC (about 0.93) rather than macro F1. Ribeiro et al. [19] address a different 12-lead abnormality task at much larger scale, and Hannun et al. [20] study single-lead ambulatory arrhythmia detection. The Results section returns to these comparisons.

## III. Methodology

### A. Data and Preprocessing

We use PTB-XL v1.0.3 [5], which holds 21,799 clinical 12-lead recordings sampled at 500 Hz, each 10 seconds long, giving tensors of shape (12, 5000). The SCP-ECG diagnostic codes on each recording map to five superclasses (CD, HYP, MI, NORM, STTC). Records whose codes mapped to no superclass were dropped, which left 21,375 recordings from 18,610 patients. NORM makes up about 43% of these records, and HYP about 6%. NORM is the normal-ECG superclass in PTB-XL, not a rhythm category. Sinus rhythm alone does not make an ECG NORM, and a record with both NORM and an abnormal code takes whichever has the higher likelihood under the rule below. HYP labels come from ECG criteria, not echocardiography, so HYP is not limited to left ventricular hypertrophy. Rhythm is a separate statement category in PTB-XL and was not used here, so HYP records were not restricted to non-sinus rhythm. No IRB approval was needed because the dataset is public, de-identified, and released under a Creative Commons license. PTB-XL was curated by the Physikalisch-Technische Bundesanstalt in Germany from recordings made with Schiller devices between 1989 and 1996, and it covers about 18,900 patients (roughly half women, ages 0 to 95, median about 62) [5]. Ethnicity is not recorded. Voltage criteria for hypertrophy differ by age, sex, and ethnic origin, and our labels come from the dataset’s annotations, so our results may not carry over to other populations.

#### Label assignment

About a quarter of the records (23.7%) carry more than one superclass, so we had to reduce each of them to a single label. We used the superclass of the highest-likelihood SCP code among codes with likelihood above zero, and we broke ties in favor of the first-listed code. For 3,130 records (14.6%) the label would change if ties went to the last-listed code. The ground truth therefore depends partly on this rule, and the per-class results carry that caveat. A single label is also a clinical limitation, because several cardiac conditions often occur together in real patients. Under our tie-breaking rule, a model is penalized in evaluation even when it correctly detects a secondary condition. This probably understates the multi-label diagnostic ability of both the deep ensembles and the classical pipeline.

No record was excluded for carrying several superclasses, and only records with no superclass code were dropped. Our F1 scores therefore measure agreement with the dominant-code label, not full ECG interpretation, and we did not measure how performance differs between records with one superclass and records with several. We did not keep the original label array, so we re-derived the labels from the released metadata. The test class counts (497, 208, 597, 1,386, and 494 for CD, HYP, MI, NORM, and STTC) fall within two records per class of those from our initial analysis, and each deep model’s test macro F1 reproduced to within 0.0011.

Each recording went through three preprocessing steps in order. A zero-phase IIR notch filter at 50 Hz (*Q* = 30) removed powerline interference, a reasonable choice for PTB-XL’s European recording setting. A zero-phase fourth-order Butterworth bandpass filter (0.5–40 Hz) removed base-line wander and high-frequency noise. Both filters used `sosfiltfilt`, so they add no phase distortion. Finally, we z-scored each lead in each recording separately, using its own mean and standard deviation within the recording, with a floor of 10*^−6^* on the standard deviation to avoid division by zero on flat or disconnected leads. Z-scoring each lead removes absolute voltage and the amplitude relationships between leads that the voltage criteria for hypertrophy depend on. The 0.5–40 Hz band also reduces QRS amplitude compared with the 150 Hz diagnostic bandwidth. Weak HYP performance may therefore reflect our preprocessing as well as class imbalance.

We did not test other normalization or bandwidth settings. The pipeline ran during signal extraction, so the stored array *X* of shape (21,375, 12, 5000) holds only preprocessed float32 signals.

We split by patient, roughly 70/15/15 into training, validation, and test sets, because a record-level split could put one patient’s several recordings on both sides. We shuffled the 18,610 patient IDs with seed 42 and divided them into disjoint groups of 13,028 training, 2,791 validation, and 2,791 test patients, giving 14,978, 3,215, and 3,182 records. The split is not stratified by class, and it differs from the recommended PTB-XL `strat_fold` partition (which also respects patient boundaries). Our absolute numbers are therefore not directly comparable with published benchmarks. The comparisons in this paper are relative, among models trained and tested under one shared split. All models share this split.

### B. Class Imbalance and Augmentation

For the deep models M1 to M4 we used two mechanisms. A `WeightedRandomSampler` weighted each training sample by the inverse of its class frequency, which oversamples minority classes within batches. We also replaced cross-entropy with focal loss [21] (*γ* = 1.0, no per-class *α* weights) and label smoothing of *ϵ* = 0.1. The focal term (1*−p_t_*)*^γ^* shifts the gradient toward hard and rare examples, and label smoothing softens the one-hot targets, which suits PTB-XL’s likelihood-based labels. Stacking all three mechanisms (the sampler, inverse-frequency *α* weighting, and focal loss with *γ* = 2.0) overcorrected in early development experiments and gave up too much NORM performance for HYP recall. So we dropped the *α* weighting and lowered *γ* to 1.0. Those runs were not kept.

We trained M6 without SMOTE [22], a choice made on validation macro F1 (0.649 without, 0.645 with), and used `class_weight=“balanced”` in LightGBM. The SMOTE variant, which expands the training set from 14,978 to 32,190 samples, is reported as an ablation.

We augmented the training data for M1 to M4 with six transforms, each applied independently with a fixed probability. Gaussian noise had a standard deviation of 1% of the signal’s own (probability 0.5). Amplitude scaling ranged from 0.9 to 1.1 (0.3), and a circular time shift reached up to *±*50 samples (0.3). One random lead was zeroed (0.15) so the model could not rely on any single lead. Simulated baseline wander, a sinusoidal drift with frequency between 0.05 and 0.5 Hz and amplitude between 0.05 and 0.15 (0.4), mimicked breathing and motion artifacts. Finally, the six limb leads (I, II, III, aVR, aVL, aVF) were randomly permuted with probability 0.2 to reduce sensitivity to lead order.

The limb-lead permutation is a problem for some labels. It changes the frontal-plane axis and moves findings such as inferior Q waves into other leads, which can change the meaning of an MI or axis-dependent conduction label. We did not run an ablation, so it may have hurt M1 to M4 on those classes.

### C. Model Architectures

#### M1 (Mamba-style SSM, pure PyTorch)

This is a selective state space model inspired by Mamba [11], written entirely in PyTorch with no custom CUDA kernels. A two-stage convolutional stem (strides 4 and 4) cuts the 5,000-sample input to 313 time steps with *d*_model_ = 128 channels. Four MambaBlock modules follow the stem. Each block applies layer normalization, an input projection to 2 *× d*_inner_ = 512 channels, a depthwise Conv1d with kernel size 4, a SiLU activation, and a selective scan with *d*_state_ = 16. We compute the scan in parallel, not as a sequential recurrence, using a prefix sum over log-space cumulative products of the discretized state transition matrices. Mean and max pooling over the 313 output steps give a 256-dimensional vector, which feeds a two-layer head with GELU activations and dropout of 0.2. The model has about 1.1 million parameters and was trained with a learning rate of 2 *×* 10*^−4^* and batch size 32.

#### M2 (multi-lead 1D residual CNN)

This ResNet-style network [8] works directly on the raw 12-lead signal of shape (12, 5000). A stem with kernel size 15, stride 2, and max pooling feeds four residual stages with 64, 128, 256, and 512 channels, each made of two ResidualBlock1D modules (kernel size 7, batch normalization, ReLU, dropout 0.2). Adaptive average pooling then reduces the time axis to a single step. A two-layer head with dropout 0.3 produces the five outputs. It has about 8.9 million parameters and was trained with a learning rate of 3 *×* 10*^−4^* and batch size 64.

#### M3 (multi-lead BiLSTM)

A bidirectional LSTM [10] runs on a convolutional summary of the raw signal. A three-stage stem with strides 4, 2, and 2 shortens the sequence from 5,000 to about 313 steps and projects to 96 channels. A two-layer bidirectional LSTM (hidden size 192, dropout 0.3) follows. Mean and max pooling over time give a 768-dimensional vector for a two-layer head. The model has about 1.5 million parameters; training used a learning rate of 5 *×* 10*^−4^* and batch size 64.

#### M4 (CRNN with attention)

A three-stage 1D convolutional extractor (kernel sizes 11, 7, and 5; strides 2, 2, and 2) produces 128 channels. A two-layer bidirectional LSTM (hidden size 256, dropout 0.3) and temporal attention come next. Attention weights come from a learned linear projection of the 512-dimensional LSTM output to a scalar, normalized by a softmax over time. The weighted sum is the final sequence representation. The model has about 2.6 million parameters; training used a learning rate of 4 *×* 10*^−4^* and batch size 64.

#### M5 (soft-voting ensemble of M1, M3, and M4, selected onvalidation)

M5 averages the softmax outputs of M1, M3, and M4 and needed no further training. Its members are the three deep models with the highest validation macro F1 (M4, M1, M3), so the selection rule uses no test information. The order of events matters here. Our first ensemble was M2+M3+M4, whose members we picked using test macro F1. We adopted the validation-based rule as the main definition only after seeing that result. We therefore report two more ensembles next to M5. M5A averages all four deep models and involves no selection. M5T is the original M2+M3+M4 ensemble. Because test results guided its choice, we treat it as exploratory and expect its advantage to be optimistic. M6 is not a neural network and serves as a separate classical baseline.

#### M6 (MiniRocket + LightGBM)

We applied the `MiniRocketMultivariate` transform from sktime [16] with 10,000 random convolutional kernels, which produces a 9,996-dimensional vector of proportions of positive values per recording. We fit the transform on a random 5,000-record subset of the *training* set only. The kernel weights do not depend on labels, but the bias values are set from the data, so fitting on training records alone keeps validation and test signals out of the transform. We then extracted features for all records in chunks of 1,000 and standardized them with a scaler fit on the training set. LightGBM [17] used at most 400 trees, `num_leaves` = 31, `learning_rate` = 0.05, `colsample_bytree` = 0.3, `subsample` = 0.8, and `class_weight=“balanced”`. Early stopping with patience 50 on the validation set chose the number of trees (221 without SMOTE, 196 with). The remaining hyperparameters were fixed in advance and not tuned.

### D. Training and Evaluation

We trained M1 to M4 with AdamW [23] (weight decay 10*^−4^*) and a cosine annealing schedule over at most 70 epochs, with early stopping (patience 12) on validation macro F1 and gradient clipping at a maximum norm of 1.0. After every epoch we saved the weights, optimizer state, best validation F1, and early-stopping counter to Google Drive so training could resume after a runtime disconnect. All deep models ran on an NVIDIA Tesla T4 in Google Colaboratory. The M6 pipeline ran on the Colab CPU runtime. The MiniRocket transform of all 21,375 records took 60.5 min (0.17 s per record), and LightGBM took 22.1 min (30.3 min with SMOTE).

Each model was trained once with seed 42, so the confidence intervals reflect sampling of the test set only, not run-to-run training variation, which we did not measure. Small gaps, such as M2 versus M3 (0.001) and M4 versus M1 (0.013), should be read with this in mind. The learning rate and batch size of each deep model were set by hand and not tuned. Training stopped through early stopping on validation macro F1 (patience 12, maximum 70 epochs), and the best-validation checkpoint was used. We did not record the epoch at which each run stopped.

Inference was rerun from each deep model’s best-validation checkpoint, with per-record class probabilities saved, and macro F1 matched the values from our initial runs to within 0.0011. All comparisons use the same 3,182 test records. Confidence intervals come from a patient-level paired bootstrap, in which we resampled test patients with replacement 10,000 times and scored every model on each resample. We also ran exact McNemar tests on per-record correctness, with Holm adjustment across all 21 model pairs among M1 to M4, M5T, M6, and M6_S_. We call two models equivalent in macro F1 only if the 90% CI of their difference lies within *±*0.02, and we report the 90% interval with every equivalence claim. We set this margin before looking at results, and it is close to the half-width of a single model’s 95% CI on this test set. When the criterion is not met, we say only that no significant difference was found. Besides macro F1, we report one-vs-rest AUROC and AUPRC (per class and macro-averaged), sensitivity, specificity, and PPV at the argmax operating point, and calibration (expected calibration error with 15 bins, Brier score, and log loss). We also report patient-level bootstrap intervals (2,000 resamples) for macro and HYP AUROC and AUPRC, including paired M6 − M5T differences. The other per-class AUROC and AUPRC values come without intervals, and we made no correction for multiple comparisons.

## IV. Results and Analysis

### A. Overall Comparison

Table II gives test performance for all models. By macro F1 the ranking is M5 (0.656), M2 (0.645), M6 (0.645), M3 (0.644), M4 (0.636), and M1 (0.624), and M6 with SMOTE scores 0.630. The all-four ensemble M5A scored 0.657, and the exploratory test-selected ensemble M5T scored 0.665. The validation-selected ensemble M5 was ahead of M1 (+0.032, 95% CI +0.020 to +0.045) and M4 (+0.019, +0.006 to +0.033), but not detectably ahead of M2 (+0.011, −0.005 to +0.027), M3 (+0.012, −0.001 to +0.025), or M6 (+0.011,−0.007 to +0.031). M5A showed the same pattern. It was ahead of M1 (+0.033, +0.020 to +0.046) and M4 (+0.020, +0.006 to +0.035), and not detectably ahead of M2 (+0.012, −0.002 to +0.026), M3 (+0.012, −0.001 to +0.026), or M6 (+0.012, −0.006 to +0.031) (Table I). We cannot call M5 and M6 equivalent, because the upper end of their 90% CI (−0.004 to +0.027) exceeds the *±*0.02 margin. Nor do our data show that soft-voting ensembles outperform MiniRocket+LightGBM on macro F1.

**TABLE 1.**
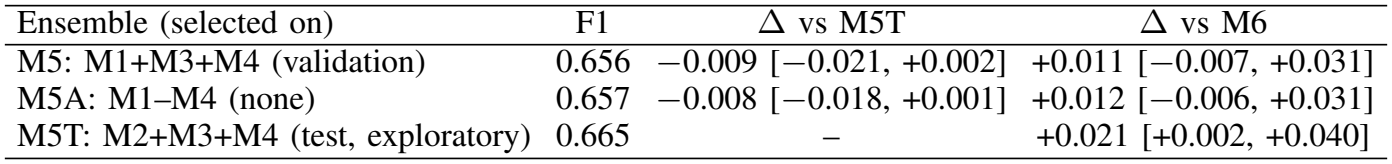
Eensembles by member-selection rule. Δ is the difference in macro F1 with its paired 95% patient-level bootstrap CI (10,000 resamples). M5 is the primary ensemble; M5T is exploratory because its members were chosen on test results.

| Ensemble (selected on) | F1 | Δ vs M5T | Δ vs M6 |
|---|---|---|---|
| M5: M1+M3+M4 (validation) | 0.656 | −0.009 [−0.021, +0.002] | +0.011 [−0.007, +0.031] |
| M5A: M1–M4 (none) | 0.657 | −0.008 [−0.018, +0.001] | +0.012 [−0.006, +0.031] |
| M5T: M2+M3+M4 (test, exploratory) | 0.665 | – | +0.021 [+0.002, +0.040] |

**TABLE 2.** Aggregate test-set performance (3,182 records). Macro F1 95% CI from a patient-level bootstrap (10,000 resamples). Precision, recall, and F1 at argmax; AUROC and AUPRC are macro one-vs-rest; ECE uses 15 bins. brier score is the multiclass version summed over the five classes, and log loss is the mean negative log-likelihood. M5 is the validation-selected ensemble (M1+M3+M4); M5T is the exploratory test-selected ensemble (M2+M3+M4). M6 is the version without smote.

| Model | Acc. | Macro F1 [95% CI] | Macro Prec. | Macro Rec. | Macro AUROC | Macro AUPRC | ECE | Brier | Log loss |
|---|---|---|---|---|---|---|---|---|---|
| M1 Mamba SSM | 0.695 | 0.624 [0.604, 0.643] | 0.617 | 0.639 | 0.889 | 0.666 | 0.054 | 0.430 | 0.833 |
| M2 1D ResNet | 0.737 | 0.645 [0.624, 0.664] | 0.652 | 0.641 | 0.906 | 0.689 | 0.026 | 0.378 | 0.735 |
| M3 BiLSTM | 0.710 | 0.644 [0.624, 0.663] | 0.637 | 0.660 | 0.905 | 0.689 | 0.027 | 0.407 | 0.772 |
| M4 CRNN+Attn | 0.715 | 0.636 [0.616, 0.656] | 0.633 | 0.642 | 0.899 | 0.683 | 0.020 | 0.395 | 0.765 |
| M5 Ensemble (M1+M3+M4) | 0.728 | 0.656 [0.636, 0.675] | 0.650 | 0.664 | 0.912 | 0.706 | 0.030 | 0.383 | 0.732 |
| M6 MiniRocket+LGBM | 0.735 | 0.645 [0.623, 0.665] | 0.676 | 0.633 | 0.914 | 0.711 | 0.038 | 0.371 | 0.715 |
| M6<sub>SMOTE</sub> | 0.724 | 0.630 [0.607, 0.651] | 0.653 | 0.617 | 0.912 | 0.702 | 0.032 | 0.377 | 0.723 |
| M5T Ensemble (M2+M3+M4), exploratory | 0.741 | 0.665 [0.644, 0.685] | 0.666 | 0.665 | 0.916 | 0.713 | 0.046 | 0.370 | 0.709 |

#### Exploratory ensemble

M5T, the ensemble of M2, M3, and M4, scored 0.665 (95% CI 0.644–0.685). Its members were chosen using test macro F1, so its margins are likely optimistic. In paired comparisons (Table III), M5T was ahead of every single deep model by 0.020 to 0.041 macro F1, with all 95% CIs excluding zero, and ahead of M6 by 0.021 (95% CI 0.002–0.040). It was the only ensemble whose paired interval over M6 excluded zero, and that interval began at only 0.002. M5 and M5A scored 0.009 and 0.008 below it (paired 95% CIs −0.021 to +0.002 and −0.018 to +0.001, both including zero). M5T scored higher than M6 in 98.5% of 10,000 bootstrap resamples, but this is a proportion and not a *p*-value. Accuracy did not differ significantly between M5T and M6 (0.741 and 0.735; McNemar *p* = 0.47 unadjusted, Holm-adjusted *p* = 1.0), and M5T’s accuracy advantage over M2 was also not significant (Holm-adjusted *p* = 1.0). Macro AUROC and AUPRC were almost the same for M5T and M6 (0.916 and 0.713 versus 0.914 and 0.711; the paired M6 −M5T differences had 95% CIs of −0.008 to +0.004 and 0.016 to +0.012).

**TABLE 3.** Paired comparisons of macro F1 (A – B) for 21 model pairs among M1 to M4, M5T, M6, and M6S. M5T is the exploratory test-selected ensemble (M2+M3+M4); paired comparisons for the validation-selected M5 are in table I and the text. the 95% CI comes from the patient-level paired bootstrap. mcnemar *p* is exact, tests per-record correctness (accuracy, not macro F1), and *p*H is the holm-adjusted value over all 21 pairs. M6S is M6 with smote. equivalence means the 90% CI lies inside ±0.02; the 95% cis shown are wider and can extend slightly past that margin for pairs marked equivalent.

| A − B | ΔF1 | 95% CI | McNemar *p*<sub>H</sub> | Equiv. |
|---|---|---|---|---|
| M1 − M2 | −0.021 | [−0.038, −0.004] | <0.001 | no |
| M1 − M3 | −0.021 | [−0.037, −0.004] | 0.329 | no |
| M1 − M4 | −0.013 | [−0.030, +0.005] | 0.083 | no |
| M1 − M5T | −0.041 | [−0.058, −0.025] | <0.001 | no |
| M1 − M6<sub>S</sub> | −0.006 | [−0.026, +0.014] | 0.003 | no |
| M1 − M6 | −0.021 | [−0.040, −0.001] | <0.001 | no |
| M2 − M3 | +0.001 | [−0.017, +0.019] | 0.001 | yes |
| M2 − M4 | +0.009 | [−0.009, +0.026] | 0.015 | no |
| M2 − M5T | −0.020 | [−0.034, −0.006] | 1.000 | no |
| M2 − M6<sub>S</sub> | +0.015 | [−0.004, +0.035] | 0.617 | no |
| M2 − M6 | +0.001 | [−0.018, +0.019] | 1.000 | yes |
| M3 − M4 | +0.008 | [−0.009, +0.024] | 1.000 | no |
| M3 − M5T | −0.021 | [−0.035, −0.007] | <0.001 | no |
| M3 − M6<sub>S</sub> | +0.014 | [−0.007, +0.036] | 0.469 | no |
| M3 − M6 | −0.000 | [−0.021, +0.020] | 0.012 | yes |
| M4 − M5T | −0.029 | [−0.042, −0.016] | <0.001 | no |
| M4 − M6<sub>S</sub> | +0.006 | [−0.014, +0.027] | 1.000 | no |
| M4 − M6 | −0.008 | [−0.029, +0.013] | 0.083 | no |
| M5T − M6<sub>S</sub> | +0.035 | [+0.016, +0.056] | 0.257 | no |
| M5T − M6 | +0.021 | [+0.002, +0.040] | 1.000 | no |
| M6<sub>S</sub> − M6 | −0.015 | [−0.027, −0.003] | 0.257 | no |

SMOTE did not help M6 in our experiments. Without it, macro F1 was higher (0.645 versus 0.630; paired difference +0.015, 95% CI 0.003–0.027), as were accuracy, AUROC, AUPRC, Brier score, and log loss (Table II). Only ECE was slightly worse.

M2 and M3 tie on macro F1 (0.645 and 0.644; paired difference 0.001, 90% CI −0.014 to +0.015, inside the equivalence margin), yet their accuracies differ (0.737 and 0.710, Holm-adjusted McNemar *p* = 0.001), so the two models trade off classes differently. The same criterion holds for M2 against M6 (+0.001, 90% CI −0.015 to +0.016) and for M3 against M6 (−0.000, 90% CI −0.017 to +0.016; its 95% interval in Table III, −0.021 to +0.020, extends just past the margin).

On macro F1, then, the CPU-only pipeline is equivalent to the single ResNet and BiLSTM (Table III). M4 scored 0.636, which is not significantly different from M2 or M3 in macro F1 (Table III). M1 scored lowest at 0.624, 0.021 below both M2 and M3 (95% CIs exclude zero); its gap to M4 (0.013) is not significant. M1 is a pure-PyTorch parallel-scan approximation of Mamba rather than the fused CUDA implementation, which may limit how well it trains. Its HYP recall (0.41) is higher than M2’s (0.32), but its HYP AUROC (0.79) is the lowest of all models, so we do not attribute any HYP advantage to SSM gating.

### Per-Class Results

The per-class tables report the single models, the validation-selected ensemble M5, M6, and the exploratory ensemble M5T. NORM, the majority class, unsurprisingly had the highest F1 for every model (0.81–0.85). F1 ranged from 0.68 to 0.71 for CD, 0.65 to 0.71 for MI, and 0.58 to 0.65 for STTC (Table IV).

**TABLE 4.** Per-class one-vs-rest AUROC and argmax F1 on the test set. M5T is the exploratory test-selected ensemble. M6 is the version without smote.

|  | AUROC |  |  |  |  | F1 (argmax) |  |  |  |  |
|---|---|---|---|---|---|---|---|---|---|---|
| Model | CD | HYP | MI | NORM | STTC | CD | HYP | MI | NORM | STTC |
| M1 Mamba SSM | 0.92 | 0.79 | 0.91 | 0.94 | 0.89 | 0.70 | 0.34 | 0.69 | 0.81 | 0.58 |
| M2 1D ResNet | 0.93 | 0.83 | 0.91 | 0.94 | 0.92 | 0.69 | 0.33 | 0.71 | 0.85 | 0.64 |
| M3 BiLSTM | 0.93 | 0.83 | 0.92 | 0.94 | 0.91 | 0.70 | 0.39 | 0.70 | 0.82 | 0.61 |
| M4 CRNN+Attn | 0.92 | 0.81 | 0.91 | 0.94 | 0.91 | 0.70 | 0.37 | 0.69 | 0.84 | 0.59 |
| M5 Ensemble (M1+M3+M4) | 0.93 | 0.84 | 0.92 | 0.95 | 0.92 | 0.71 | 0.40 | 0.71 | 0.84 | 0.62 |
| M6 MiniRocket+LGBM | 0.92 | 0.87 | 0.91 | 0.94 | 0.92 | 0.70 | 0.37 | 0.65 | 0.85 | 0.65 |
| M6<sub>SMOTE</sub> | 0.92 | 0.87 | 0.90 | 0.94 | 0.92 | 0.68 | 0.35 | 0.65 | 0.85 | 0.63 |
| M5T Ensemble (M2+M3+M4), exploratory | 0.94 | 0.85 | 0.92 | 0.95 | 0.92 | 0.71 | 0.41 | 0.71 | 0.85 | 0.64 |

**Fig 1.**
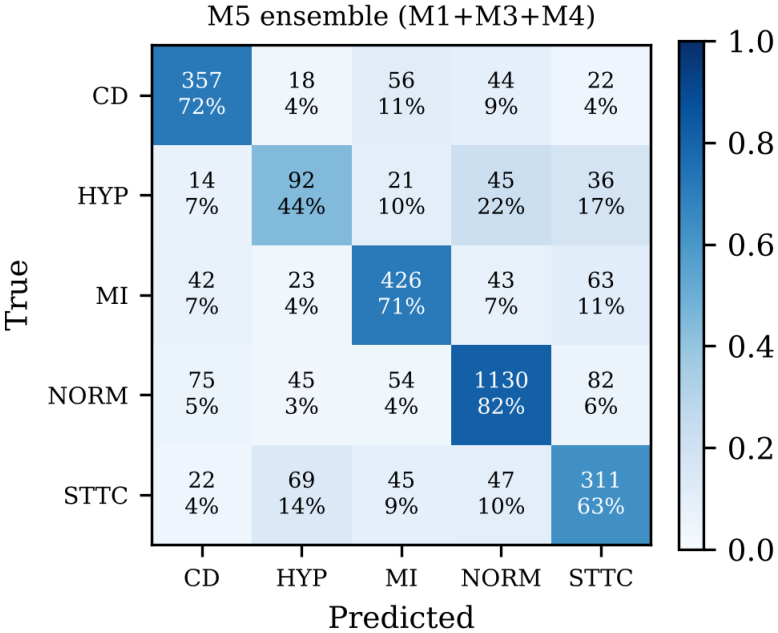

Confusion matrix for the validation-selected ensemble M5 (M1+M3+M4) on the test set. Cells show counts and row percentages.

**Fig 2.**
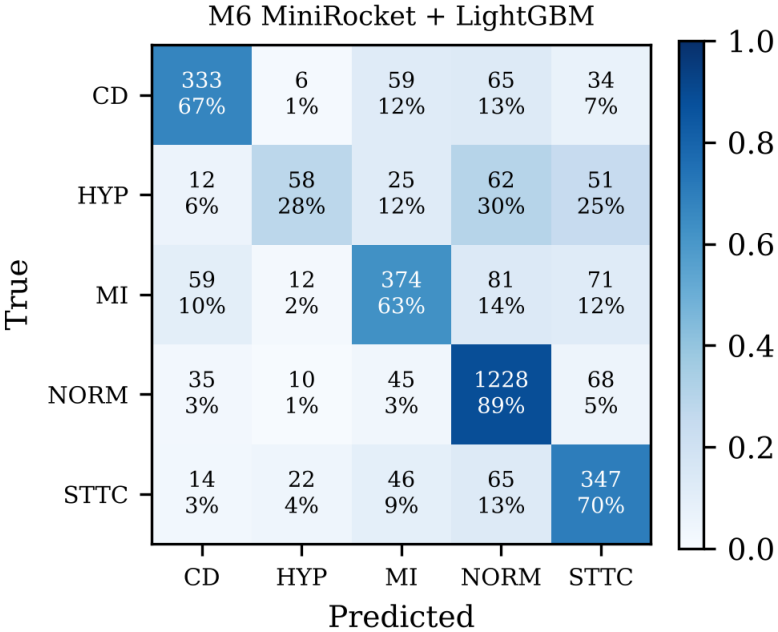

Confusion matrix for M6 (MiniRocket + LightGBM, no SMOTE) on the test set. Cells show counts and row percentages.

**TABLE 5.** Hypertrophy (HYP) detail on the test set (208 records). Sensitivity, specificity, PPV, and F1 are at argmax. M5T is the exploratory test-selected ensemble. M6 is the version without smote.

| Model | AUROC | AUPRC | Sens. | Spec. | PPV | F1 |
|---|---|---|---|---|---|---|
| M1 Mamba | 0.791 | 0.284 | 0.409 | 0.928 | 0.285 | 0.336 |
| M2 ResNet | 0.825 | 0.309 | 0.317 | 0.957 | 0.340 | 0.328 |
| M3 BiLSTM | 0.830 | 0.304 | 0.490 | 0.929 | 0.327 | 0.392 |
| M4 CRNN | 0.813 | 0.308 | 0.423 | 0.940 | 0.331 | 0.371 |
| M5 Ensemble (M1+M3+M4) | 0.839 | 0.342 | 0.442 | 0.948 | 0.372 | 0.404 |
| M6 MiniRocket | 0.871 | 0.423 | 0.279 | 0.983 | 0.537 | 0.367 |
| M6 (SMOTE) | 0.871 | 0.403 | 0.284 | 0.975 | 0.444 | 0.346 |
| M5T Ensemble (exploratory) | 0.849 | 0.355 | 0.413 | 0.959 | 0.413 | 0.413 |

HYP was the weakest class for every model by a wide margin (F1 0.33–0.41, Table V). It makes up about 6% of records (208 of the 3,182 test records). Differences in HYP recall at argmax mostly reflect where each model sits on the sensitivity/PPV trade-off, not better features. M3 has the highest recall (0.49) at a PPV of 0.33, while M6 has the lowest (0.28) at a PPV of 0.54. M3’s higher recall reflects a lower PPV rather than a better ranking, because its HYP AUROC (0.830) is below those of M6 (0.871), M5T (0.849), and M5 (0.839). This fits a different operating point and not better features. HYP labels come in part from voltage criteria (atrial overload and enlargement, also grouped under HYP, are not voltage-based), and our preprocessing removes absolute and inter-lead amplitude. Before any threshold is applied, M6 has the highest HYP AUROC (0.871, 95% CI 0.846–0.897) and AUPRC (0.423, 0.353–0.498), ahead of M5 (0.839; 0.342) and M5T (0.849, 0.822–0.875; 0.355, 0.289–0.427). The paired M6 − M5T differences were +0.001 to +0.043 for AUROC (the lower bound is barely above zero) and +0.022 to +0.114 for AUPRC. These results say little about what architectures can learn about hypertrophy.

Counting false-normal predictions under our single-label rule, M5 labeled 179 of 1,796 abnormal test records (CD, HYP, MI, or STTC; 10.0%) as NORM, M5T labeled 215 (12.0%), and M6 labeled 273 (15.2%). The paired M6 − M5 difference was +0.052 (95% CI +0.039 to +0.066), and the M6 − M5T difference was +0.032 (95% CI +0.019 to +0.046).

### C. Calibration and Cost

Calibration varied more across models than macro F1 did (Table II, Fig. 3). M4 had the lowest ECE (0.020) and M1 the highest (0.054), with M1 overconfident at high confidence. The exploratory ensemble M5T had the best Brier score (0.370) and log loss (0.709), narrowly ahead of M6 (0.371 and 0.715), but its ECE was 0.046. The primary ensemble M5 had an ECE of 0.030, a Brier score of 0.383, and a log loss of 0.732. These are comparable to the ResNet (0.026, 0.378, and 0.735) and better than M1, M3, and M4 on Brier score and log loss, although its ECE is worse than that of M2, M3, and M4. Both ensembles are underconfident on average (mean confidence 0.698 for M5 and 0.694 for M5T, against accuracies of 0.728 and 0.741), which is common when probabilities are averaged across models. No model was calibrated after training. Macro F1 at argmax depends on the threshold and on class prevalence, so it says little about whether a model is ready for deployment. Table VI lists the multiply-accumulate counts (MACs) from the `thop` library for one forward pass on a (1, 12, 5000) input. FLOPs are roughly twice these values. We estimated M1 by hand, since thop cannot profile its parallel cumulative-sum scan, so that value may not be directly comparable. Among the deep models, M2 is the most expensive at about 3.95 billion MACs and 8.9 million parameters, because its four-stage convolutional stack runs on long sequences. M3 needs about 951 million MACs with 1.5 million parameters, and M4 about 3.26 billion with 2.6 million. The cost and training time of each ensemble are simply the sums of its members. M5 (M1+M3+M4) needs about 4.9 billion MACs, 5.2 million parameters, and about 8.5 h on a T4, while M5T (M2+M3+M4) needs about 8.2 billion MACs and about 9.3 h.

**TABLE 6.**
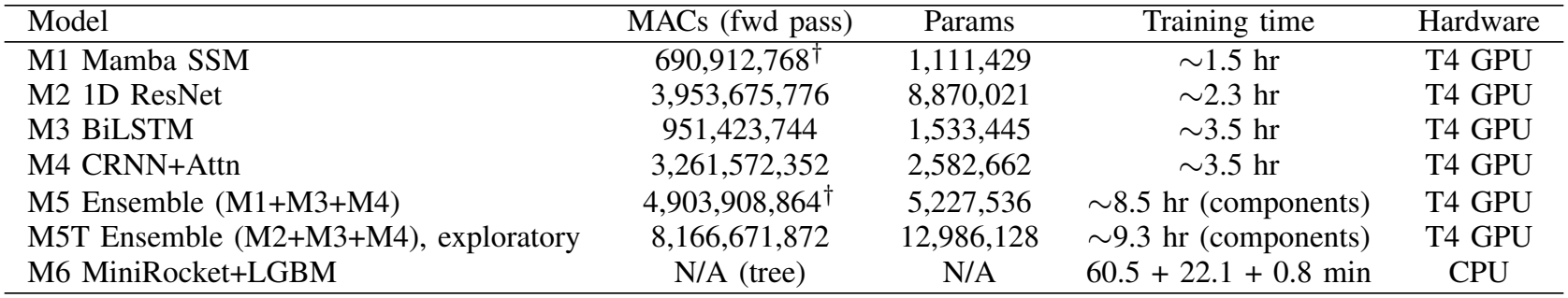
Computational cost. MACs are for one forward pass as counted by `thop`. The M1 value (†) is an analytic estimate, so ensemble macs that include M1 (†) inherit that caveat. Each ensemble’s training time is the sum of its components. T6 is the version without SMOTE; its training time is the minirocket transform, the lightGBM fit, and the 0.8 min transform fit.

**Fig 3.**
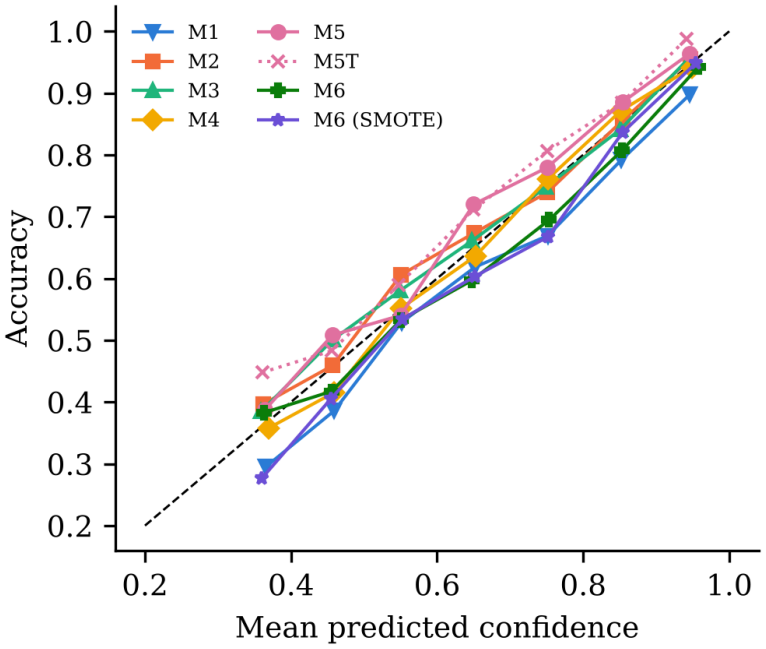

Reliability diagram on the test set (bins with fewer than 20 records omitted). M5 is the validation-selected ensemble (M1+M3+M4) and M5T (dotted) is the exploratory test-selected ensemble. M6 is the version without SMOTE.

M6 trained in about 83 min on CPU (60.5 min for the MiniRocket transform, 22.1 min for LightGBM; 92 min with SMOTE, both including the 0.8 min transform fit), about the same as M1’s roughly 90 min on a T4, so we do not claim a large wall-clock saving. A GPU is usually needed to train neural networks and is not routinely available in many clinical settings, whereas a CPU is a general-purpose processor found in any computer. M6 needs no GPU, but inference is not free. The MiniRocket transform alone took 61 minutes across the whole dataset, a figure that depends on hardware, and we did not time inference for the deep models or the final classifier. M1 has 1.11M parameters, close to M3’s 1.53M, yet scored below both M3 and M2 (8.87M), so we make no claim that it is parameter-efficient. We did not test either approach in a clinical workflow.

### D. Comparison with Published Literature

Strodthoff et al. [9] frame the superdiagnostic task as a multi-label problem and report a macro AUC of about 0.93 for their best models, whereas we use single-label macro F1 on a custom split (our one-vs-rest macro AUROC is 0.89–0.92). Ribeiro et al. [19] and Hannun et al. [20] study different tasks (12-lead abnormality detection and single-lead ambulatory arrhythmia detection) on much larger datasets. Because our split is patient-disjoint but not the recommended strat_fold partition, we cannot attribute any gap with published numbers to stricter evaluation.

## V. Conclusion and Discussion

We compared six ECG classification approaches on 21,375 PTB-XL records using one patient-level split throughout. Our main finding is that no model clearly outperformed MiniRocket+LightGBM on macro F1. The ensemble selected on validation (M5, M1+M3+M4) scored 0.656, and the all-four ensemble scored 0.657. Both were ahead of the Mamba-style and CRNN models, but neither was detectably ahead of the ResNet, the BiLSTM, or MiniRocket+LightGBM (+0.011 and +0.012 macro F1 over M6, with 95% CIs including zero). The exploratory ensemble of M2, M3, and M4 reached 0.665 and was modestly ahead of MiniRocket+LightGBM (+0.021, 95% CI 0.002–0.040), but its members were chosen on test results, so we treat that margin as optimistic. MiniRocket+LightGBM came close on accuracy and threshold-free metrics and had a higher HYP AUROC than both ensembles (0.871 versus 0.839 for M5 and 0.849 for M5T), although it labeled more abnormal ECGs as NORM (15.2% versus 10.0% for M5 and 12.0% for M5T). Under our *±*0.02 criterion it was also equivalent to the ResNet and BiLSTM on macro F1. It also trains without a GPU and did not benefit from SMOTE. The Mamba-style model scored below the CNN and BiLSTM. Differences in HYP recall mostly reflect threshold choice, and our preprocessing, which removes amplitude information, may also limit HYP performance.

The dataset is fairly small, and the conclusions could change with a larger *N* . We used only one dataset and one split. We had no external validation, subgroup analysis, or clinically defined operating points, and we did not evaluate on the recommended `strat_fold` partition, so our absolute numbers cannot be compared directly with published benchmarks. None of the models used self-supervised pretraining, which could change the ranking, and we did not test a 1D vision transformer. We adopted the validation-based ensemble rule after first examining the test-selected ensemble, which is why we report that ensemble as exploratory. The single-label ground truth depends on a tie-breaking rule that changes 14.6% of labels, and 23.7% of records carry more than one superclass. Z-score normalization, the 40 Hz low-pass filter, and limblead permutation may have hurt HYP, MI, and axis-dependent classes, and we did not ablate them. The equivalence margin was our own choice. Each model was trained once, so our intervals do not capture training variability. Intervals for AUROC and AUPRC cover only macro and HYP values, use 2,000 resamples, and are not corrected for multiple comparisons, and no model was calibrated.

Future work includes pretraining, evaluation on the official `strat_fold` partition (fold 9 for validation, fold 10 for testing) with repeated cross-validation and multiple training seeds, so that results can be compared with published benchmarks, multi-label evaluation, ablations of preprocessing and augmentation, and external clinical validation before any deployment.

## VI. Data and Code Availability

The PTB-XL dataset is available at https://physionet.org/content/ptb-xl/. Code for model training, evaluation, and boot-strap analysis, together with the patient and record order, the split indices, the re-derived single-label array and the rule that produced it, and the library versions used for the analysis, is available at https://github.com/Vivaan-bhatt/ecg-project and archived on Zenodo (version v1.0, https://doi.org/10.5281/zenodo.24148794).

